# Albuminuria Changes as a surrogate endpoint in *Apolipoprotein L1* Mediated Kidney Disease in Vanderbilt BioVU and the Million Veteran Program

**DOI:** 10.64898/2026.06.04.26354945

**Authors:** Fatih Mamak, Zhihong Yu, Jefferson L. Triozzi, Robert Corty, Lee Wheless, Guanchao Wang, Ayush Giri, Hua Chang Chen, Otis Wilson, Alexander G. Bick, J. Michael Gaziano, Ran Tao, Adriana M. Hung

## Abstract

**Importance:** Recently, proteinuria has been accepted as a surrogate end point for clinical trials in focal segmental glomerulosclerosis (FSGS) ang IgA nephropathy. However, proteinuria has not been evaluated in Apolipoprotein L1 (APOL1)-mediated kidney disease (AMKD).

**Methods:** Real world data (RWD) analysis of 128 patients of African ancestry with APOL1 high risk genotypes, without diabetes, enrolled in the Million Veteran Program (MVP; n=109) or the biorepository at Vanderbilt University (BioVU; n=19), who had urine albumin-creatinine ratio (UACR) >= 420 mg/g (PCR∼0.9 g/g) with a concurrent GFR value. The main predictor was change in the log-UACR at 12 months. The primary outcome was annual GFR slope over 24 months. Secondary outcomes included a kidney composite of a sustained 30% GFR decline, end stage kidney disease (ESKD) or death and ESKD as a single outcome. Linear regression and Cox proportional hazards models were used to assess the effect of changes in UACR and the outcomes.

**Results:** In the pooled analysis the mean age was 56.8 (SD 15.5) y, 116 were male (90.6%) and three patients had diagnosis of FSGS at baseline. Mean baseline eGFR was 46.8 (SD 16.1) mL/min/1.73m2, mean baseline UACR was 1240.8 (1107.7) mg/g, mean eGFR slope was - 4.67[-6.00, -3.33] mL/min/1.73m2/year and the geometric mean percentage changes in the UACR at 12 months were -57.5% [-65.0%, -48.4%]. For every 1 unit of log (UACR) increment at 12 months, the annual eGFR slope decreased by -1.80 [-2.56, -1.03] mL/min/1.73m^2^ in the pooled analysis. For every 1 unit of log (UACR) increment at 12 months, the Cox regression showed a 61% increase in the risk of a kidney composite (p=0.002) and a 98% increase in the risk of ESKD (p<0.001). It was estimated that a 50% reduction of UACR at 12 months was associated with a 28% reduction in the kidney composite endpoint (adjusted hazard ratio [aHR]=0.72; 95% confidence interval [CI]:0.59-0.88; p=0.002), and a 38% reduction in the risk of ESKD (aHR=0.62; 95% CI:0.49-0.80; p<0.001).

**Conclusions and relevance:** Changes in UACR at 12 months significantly modify the rate of decline of GFR over 24 months and clinically meaningful endpoints, supporting the use of UACR changes as surrogate endpoint in AMKD.

## Introduction

African Americans have a four-fold increase in the risk of progressing to end stage kidney disease (ESKD), which has been partially explained by Apolipoprotein L1 (APOL1) high risk variants G1 and G2^1,2^. The prevalence of the high-risk genotype (2 copies of the risk alleles) is 12-15% in African Americans in different cohorts^3,4^. Although multiple mechanisms have been proposed for the nephrotoxicity of G1 and G2, APOL1 mediated kidney disease (AMKD) is recognized predominantly as a podocytopathy ^5,6^. The phenotype of AMKD is heterogenous, spanning from FSGS (all varieties) with nephrotic syndrome, to patients with diffuse or global glomerulosclerosis with sub-nephrotic range or no proteinuria often considered hypertensive kidney disease or arterionephrosclerosis, but also including other kidney manifestations that potentially can experience fast progression depending on different factors such as the inflammatory response involved^2,7^. However, in general those with proteinuria have faster progression and in them proteinuria is considered a marker of ongoing podocyte injury and disease progression^2,8^.

Proteinuria reduction has been associated with decreased risk of progression in several forms of CKD. Recent studies of focal segmental glomerulosclerosis (FSGS) have shown that proteinuria reduction short of complete remission are strongly correlated with renal survival^9^. In FSGS cohorts, despite its heterogeneity, they showed that proteinuria reduction to <0.7 g/g, over a two year period was associated with 85% (Parasol) and 92% (RaDaR) reduction in the risk of kidney failure risk over the next 5 years^10^. However, eGFR slope endpoints had a much weaker association, being unfeasible to attain an eGFR endpoint for FSGS in these studies.

Another recent study in IgA nephropathy demonstrated that both proteinuria (6-24 months) and eGFR slope (6-30 months) strongly associated with kidney failure over 5 years^11^. Based on these findings, proteinuria has become an accepted surrogate endpoint for clinical trials in IgA^11,12^.

Surrogate end points for the clinical outcome of kidney failure have been accepted by the US Food and Drug Administration (FDA). In March 2018, the National Kidney Foundation, FDA, and European Medicines Agency (EMA) jointly organized a conference focused on establishing surrogate end points for kidney failure. It was concluded that GFR decline >30%-40% and GFR slope between-arm reduction >0.5-1.0 mL/min/1.73m^2^ per year were valid surrogate end points for kidney failure^13^. It was considered that changes in albuminuria may be informative when baseline albuminuria is large and has a dominant role in disease progression and the therapeutic under investigation has a mechanistic relationship to reduce albuminuria. Hence in these specific circumstances, urinary albumin-creatinine ratio (UACR) reduction >30% was deemed a reasonably likely surrogate end point for kidney failure in some trials and valid surrogate end points in others^13^

Rosenberg et al. used individuals from the AASK, CRIC and ARIC, and generated surrogate end-points at 3 years, which included 3-year 30% and 40% decline in GFR, >3 mL/min/1.73m^2^ per year decline in GFR and doubling of the urine protein to creatinine ratio (UPCR). Individuals with APOL1 high risk genotype, had an increased risk of all 3 GFR outcomes. In the analysis of AASK and CRIC, patients with high-risk genotype with doubling of UPCR from baseline to three years had four times increased risk of kidney failure. This study suggests the potential of using surrogate markers for AMKD, which are particularly relevant in light of ongoing trials^14^.

Given that in AMKD proteinuria plays a key role in disease progression, we evaluate if changes in UACR at 12 months in MVP and BioVU are associated with annual GFR slope at 24 months and more importantly with decreased risk of clinically meaningful endpoints, such as sustained 30% decline in eGFR, ESKD or death in patients with AMKD. Addressing this question will add to the body of knowledge of ongoing studies that aim to inform if changes in UACR can represent a surrogate outcome and predict treatment effect on patient outcomes. This could accelerate drug development and approval for treatments that significantly impact patient outcomes^15^.

## Methods

### Study Population and Design

We assembled a retrospective cohort of individuals of African ancestry with 2 APOL1 high risk variants (G1 or G2), and UACR ≥ 420 mg/g (∼0.9 g/g UPCR) without diabetes. Participants were identified from two biobanks, the VA Million Veteran Program (MVP)^16^ and the Vanderbilt University Medical Center’s, EMR-linked biobank (BioVU)^17^ to evaluate if changes in UACR at 12 months could predict kidney outcomes.

The VA MVP is a VA national cohort launched in 2011 designed to study the contributions of genetics, lifestyle, and military exposures to health and disease among US Veterans^16,18^ . BioVU is the Vanderbilt University Medical Center’s biobank launched in 2007 linking de-identified EMR data (synthetic derivative) to genetic information, housing genetic data for over 360,000 participants with DNA available^17,19^ ^20,21^. Both biobanks offer a wealth of phenotypic information and longitudinal EMR data spanning over at least two decades.

The date of cohort entry for the study, was the earliest^22^ occurrence of UACR ≥ 420 mg/g with a qualifying eGFR value (≥25 and <90 mL/min/1.73 m²) within a window spanning 60 days prior to 120 days following the qualifying UACR. The overall study design is illustrated in **Supplemental Figure 1**. The UACR on the index date and the nearest qualifying eGFR measurement were designated as baseline values. The main exposure of interest was UACR change at 12 months (with a ±61-day window). The primary outcome was the annual eGFR slope over 24 months.

The secondary outcome included a time to a kidney composite endpoint of a sustained ≥30% decline in eGFR, ESKD or death and ESKD as a single endpoint. Both studies were IRB approved by their correspondent IRB, the VA Central institutional review board (IRB) and the Vanderbilt IRB under exempt categories.

### Inclusion and exclusion criteria

Patients in MVP or BioVU were eligible for inclusion in the initial study cohort if they met all of the following criteria: (1) age ≥18 years; (2) presence of two APOL1 risk variants; (3) at least one UACR value ≥420 mg/g; and (4) at least one qualifying eGFR value (≥25 and <90 mL/min/1.73 m²) documented within 60 days before to 120 days after the first occurrence of UACR ≥420 mg/g. Fulfillment of criteria (3) and (4) defined the index date, which served as the baseline time point for the study and the basis for a series of exclusion criteria.

Exclusion criteria included: diagnosis of any of the following diseases prior to or within 90 days after the index date: diabetes type 1 or 2, HIV, lupus erythematosus, autosomal dominant polycystic kidney disease (ADPKD), Alport syndrome, sickle cell disease, liver cirrhosis, or ESKD; or lack of follow-up UACR (∼12 months) or eGFR values (24 months) after the index date. Patients eligible for the analysis are shown in **Figure 1**.

**Figure 1.**
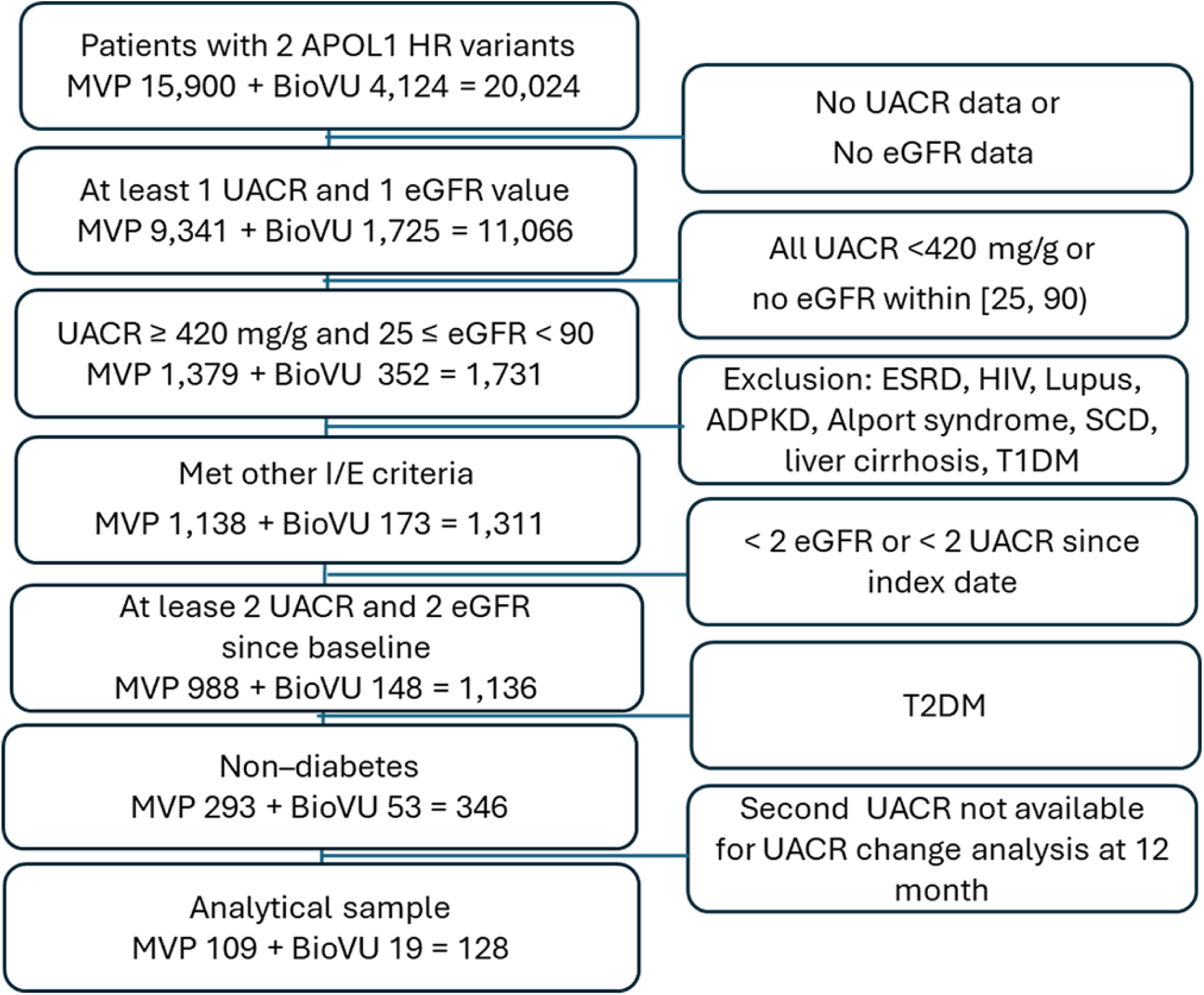
Patient flow chart for primary analysis

#### Main exposure

The primary exposure was proteinuria change at 12 months after the index date. Proteinuria was assessed using the UACR, which was extracted directly from the EMR when available, or derived from the available UPCR using the formula developed by the CKD Prognosis Consortium^22^. UACR change at 12 months was calculated as the natural logarithm of the ratio of follow-up UACR to baseline UACR. The 1-unit decrement or increment of log (UACR) corresponds to 63% decrease or 2.72-fold change at UACR scale. To maximize data capture, a ±61-day window around the 12-month time point was applied; if multiple UACR values were available within this window, the measurement closest to the exact 12 months’ time point was used.

#### Study outcomes and follow up

The primary outcome was the annual slope of eGFR that was estimated by a linear mixed-effects model fitted to longitudinal eGFR data from the index date through 24 months. The secondary outcomes were the clinical meaningful endpoints of 1) a kidney composite outcome of a sustained ≥30% decline in eGFR from baseline (confirmed by ≥2 qualifying declines ≥28 days apart), ESKD or death; 2) ESKD as a single outcome.

#### APOL1 Genotype

APOL1 high-risk variants G1 (rs73885319 p.S342G; rs60910145 p.I384M) and G2 (rs71785313, a 6–base pair deletion that removes amino acids N388 and Y389), were directly genotyped on DNA that was extracted from whole blood in both cohorts^4,23^. Participants were defined as high risk if they had 2 high risk alleles: homozygotes for G1/G1, homozygotes for G2/G2, or compound heterozygotes for G1/G2. All genotypes were in Hardy-Weinberg equilibrium and present at frequencies comparable to prior reports and in population databases^4,23^.

#### Covariates

Characteristics reported include demographics, comorbidities, laboratory tests, and the use of renin angiotensin aldosterone system inhibitors (RAASi) or sodium-glucose co-transporter-2 inhibitors (SGLT2i). Creatinine was used to calculate eGFR using the race free Chronic Kidney Disease Epidemiology Collaboration equation ^24^. In this study all individuals were of African ancestry. Ancestry determination was genetically inferred using global ancestry estimation. Admixture proportions were inferred by the Alliance of Genetic Discovery/BioVU using SCOPE, which estimates an individual allele frequency matrix through latent subspace estimation and then decomposes the estimated matrix into ancestral allele frequencies and admixture proportions^20,25^ . For MVP we also used genetically inferred ancestry informed by global ancestry using a similar approach which was generated by the MVP genetic core^26^.

## Statistical analysis

Descriptive statistics were used to summarize baseline characteristics, with frequencies and percentages reported for categorical variables and means with standard deviations for continuous variables. The annual slope of eGFR change was estimated using a linear mixed-effects model fitted to longitudinal eGFR data, treating follow-up time (in years) as a fixed effect and including both random intercepts and random slopes with patient ID as the clustering variable. Individual annual slopes of eGFR were extracted from the fitted model. Linear regression models were used to evaluate the association between UACR change at 12 months and annual eGFR slope. Cox proportional hazards models were applied to assess the effect of UACR change at 12 months on the kidney composite endpoint and ESKD. Identical analyses were performed separately in the MVP and BioVU cohorts, and results were combined using fixed-effect meta-analysis. The decision to meta-analyze both datasets was to increase generalizability by increasing the number of female and younger participants and to increase sample size. Sequential models were used for both the linear and the Cox regression models. Model 1 was adjusted for baseline log transformed UACR, Model 2: adjusted for covariate in Model 1 and age and sex; and Model 3: adjusted for covariates in Model 2 and the use of ACE inhibitors or angiotensin receptor blockers during the year prior to the index date. All analyses were conducted in *R version 4.2.2* , and the packages *lme4*, *survival* , and *mvmeta* ^27^ were used for building mixed-effect models, Cox models, and meta-analysis.

## Sensitivity analysis

A sensitivity analysis was conducted using a lower GFR threshold of 75 ml/min commonly used in CKD clinical trials^28^ and at lower UACR threshold of 321 mg/g (∼0.7 g/g UPCR). These threshold were explored based on the threshold reported for FSGS in PARASOL and RaDar below which the risk of clinically meaningful outcomes reduced significantly. ^10^

## Results

### Baseline and Clinical Characteristics for the Participants

Among 4,124 patients with two APOL1 high-risk variants identified in BioVU, the index date (as defined in supplemental Figure 1) could be determined for 352 (8.5%). After applying all exclusion criteria, 53 patients remained in the final study cohort. Of these, UACR change at 12 months could be calculated for 19 patients. Among 15,900 patients with two APOL1 high-risk variants identified in MVP, the index date could be determined for 1,379 (8.7%). After applying all exclusion criteria, 293 patients remained in the final study cohort. Of these, UACR change at 12 months could be calculated for 109 patients. The final analytic dataset included 128 patients with APOL1 HR genotypes and UACR values ≥420 mg/g at baseline (**Figure 1**).

In the study cohort the mean age was 56.8 (SD 15.5) y, 116 were male (90.6%) and three patients from BioVU had diagnosis of FSGS at baseline. The 128 patients represented patients with UACR close to 12 months and longitudinal eGFR data for the assessment. Baseline eGFR was 46.8 (16.1) mL/min/1.73m^2^ and it was similar in both studies, baseline UACR was 1240.8 (1107.7) mg/g, 91 (71.1%) of the patients were on ACE inhibitors or angiotensin receptor blockers as expected and no patients were on sodium-glucose transport 2 (SGLT2) inhibitors for this cohort. The mean (SD) time of follow up was 3.7 (3.8) y in MVP and 3.1 (3.9) y in BioVU. In MVP, while no patients had FSGS recorded at baseline, 7 patients developed FSGS during the follow up time based on an ICD-10 code plus CPT code for a kidney biopsy and were assumed to have FSGS (**Table 1**).

**Table 1.** Clinical characteristics for cohort in primary analysis (i.e., baseline UACR ≥ 420 mg/g; 25 ≤ baseline eGFR< 90)

|  | **BioVU** | **MVP** | **Pooled** |
| --- | --- | --- | --- |
| No. patients | 19 | 109 | 128 |
| Age, years (mean, SD) | 46.2 (16.4) | 58.6 (14.6) | 56.8 (15.5) |
| Males (n, %) | 13 (68.4%) | 103 (94.5%) | 116 (90.6%) |
| eGFR, mL/min/1.73m<sup>2</sup> (mean, SD) | 49.2 (18.0) | 46.4 (15.8) | 46.8 (16.1) |
| UACR, mg/g (mean, SD) | 1799.3 (1389.3) | 1143.4 (1028.0) | 1240.8 (1107.7) |
| ACEi or ARBs (n, %) | 9 (47.4%) | 82 (75.2%) | 91 (71.1%) |
| SGLT2 (n, %) | 0 (0%) | 0 (0%) | 0 (0.0%) |
| FSGS, baseline (n, %) | 3 (15.8%) | 0 (0%) | 3 (2.3%) |
| FSGS, within follow up (n, %) | 3 (15.8%) | 7 (6.4%) | 10 (7.8%) |
| Follow-up, yrs (mean, SD) | 3.1 (3.9) | 3.7 (3.8) | 3.6 (3.8) |
| Follow-up, yrs (median, IQR) | 1.4 [0.9;4.8] | 2.6 [1.1; 4.5] | N/A* |
| Annual eGFR slope over 24 months (mL/min/1.73m<sup>2</sup>/year), 95% CI | -11.82<br>[-16.50, -7.15] | -4.03<br>[-5.42, -2.64] | -4.67<br>[-6.00, -3.33] |
| Geometric mean percentage change in proteinuria at 12 months (% , 95% CI) | -56.3%<br>[-76.9%, -17.4%] | -57.6%<br>[-65.6%, -47.9%] | -57.5%<br>[-65.0%, -48.4%] |
| ***Meaningful clinical endpoints*** |  |  |  |
| 30% decline in eGFR | 12 (63.2%) | 71 (65.1%) | 83 (64.8%) |
| End stage kidney disease | 10 (52.6%) | 49 (45.0%) | 59 (46.1%) |
| death | 1 (5.3%) | 34 (32.1%) | 35 (27.3%) |
| Composite | 13 (68.4%) | 81 (74.3%) | 94 (73.4%) |

N/A\*: Not Available, since there is no feasible approach to get pooled median IQR without combining individual data.

### Primary outcome: Association between UACR change at 12 months and annual slope of eGFR over 24 months

For patients with APOL1 mediated kidney disease, we were able to demonstrate an inverse association of UACR changes at 12 months and the annual eGFR slope over 24 months. In the pooled analysis, for every 1 unit of log (UACR) increment, the annual eGFR slope decreased in Model 3 (adjusted for baseline log UACR, age, sex, and use of RAASi by -1.80 [-2.56, -1.03] mL/min/1.73m^2^/year (p<0.001). These effects were statistically significant in the pooled and MVP but not significant in BioVU (**Table 2**).

**Table 2.**
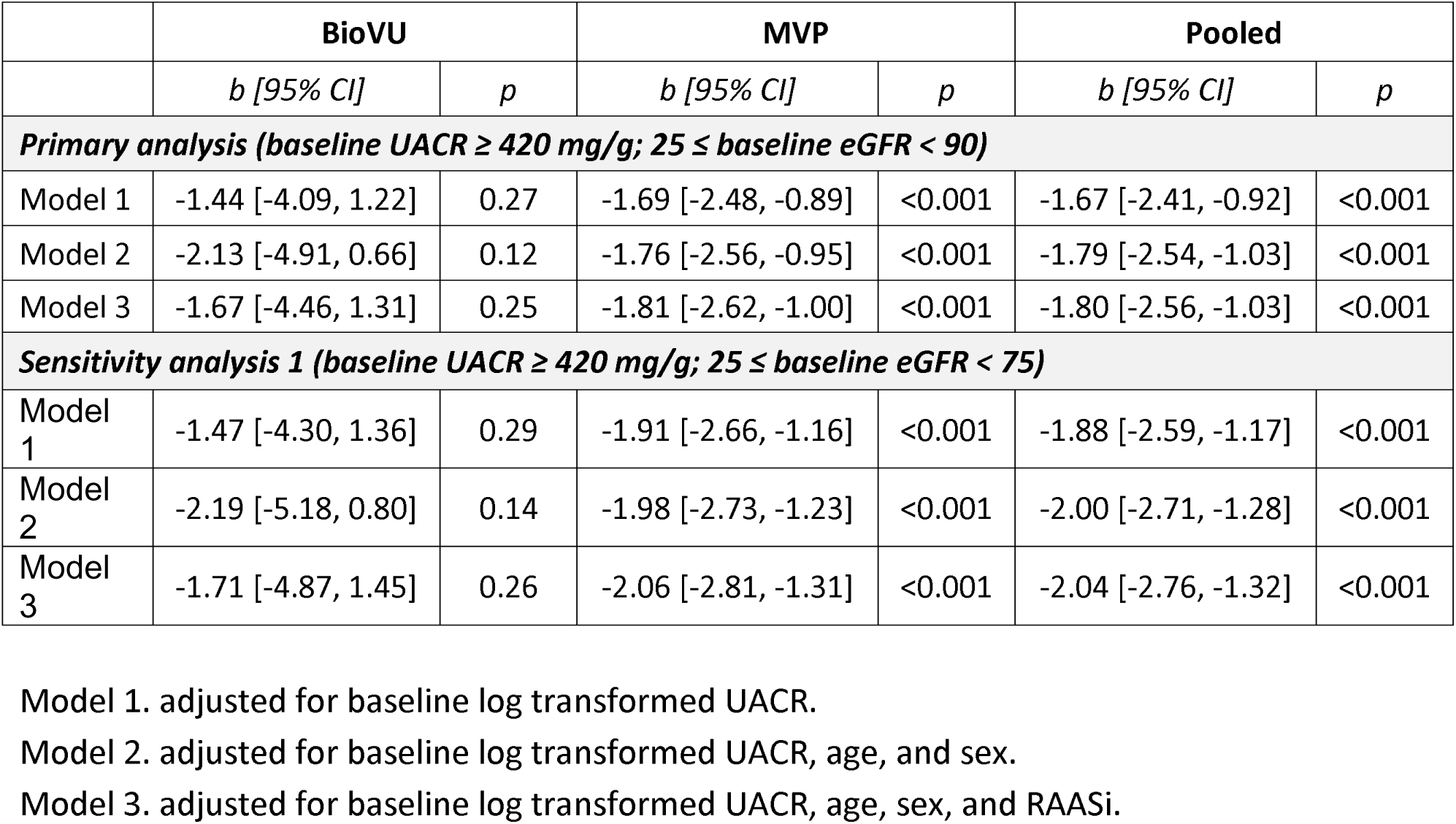
Association between log-UACR changes from baseline to 12 months and eGFR slope over 24 months among non-diabetic patients with 2 APOL1 high risk variants with proteinuria.

We predicted the effect of reductions of UACR changes at 12 months on the annual eGFR slope over 24 months based on the pooled Model 3. For example, when holding all adjustments constant, persistent proteinuria didn’t lead to any change of annual eGFR slope, while a 30%, 50%, and 90% UACR reduction increased the annual eGFR slope by 0.64 [0.37, 0.91], 1.25 [0.71, 1.78], and 4.14 [2.37, 5.90] mL/min/1.73m^2^/year, respectively (**Figure 2**).

**Figure 2.**
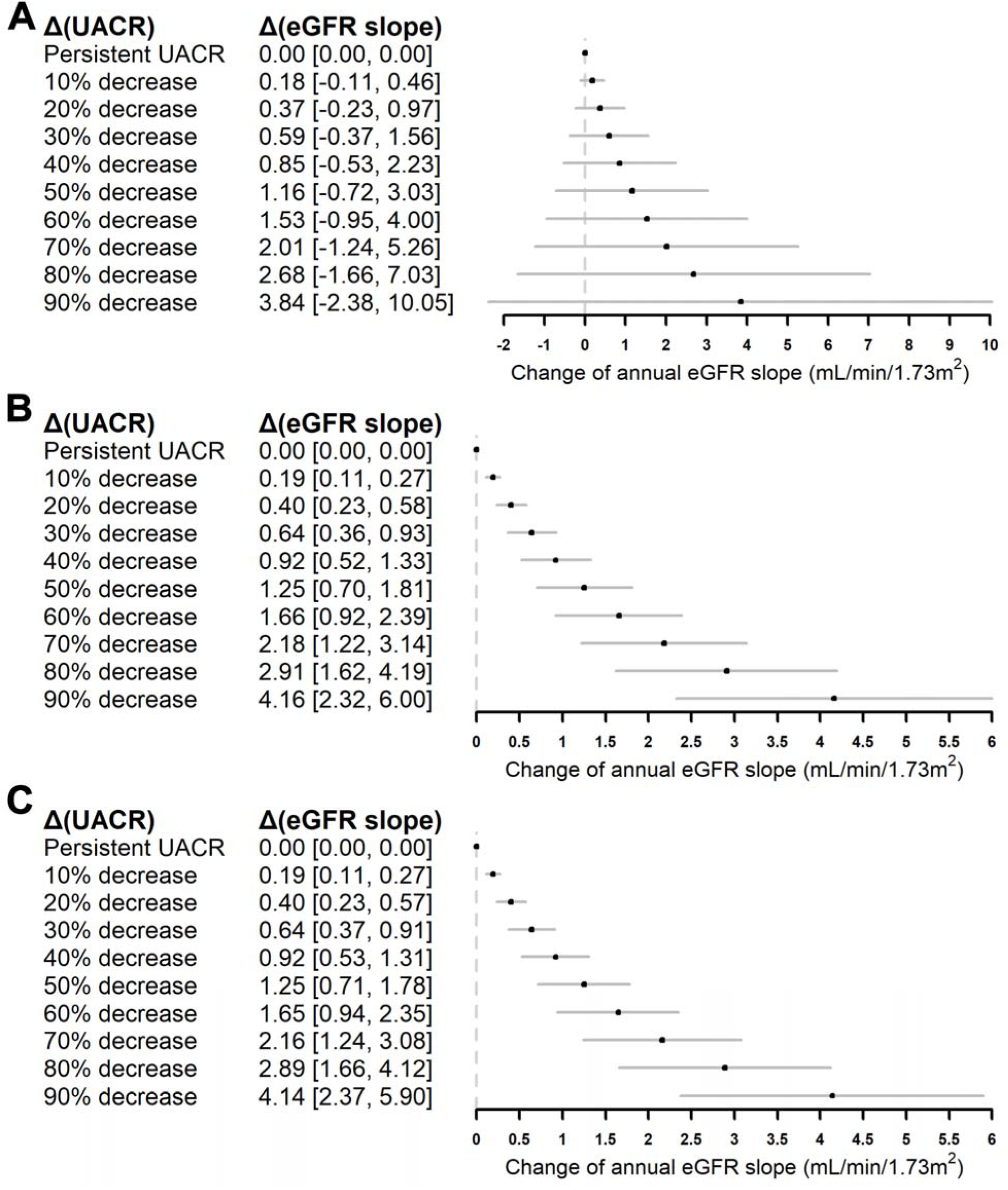
The predicted effect of 12 months UACR changes on the change of annual eGFR slope over 24 months from the Model 3 for BioVU (A), MVP (B), and Pooled cohort (C).

### The effect of UACR change at 12 months on the composite clinical outcome of 30% decline in eGFR, ESKD or death

The median follow up time was 1.4 [0.9;4.8] and 2.6[1.1; 4.5] years in BioVU and MVP. During the follow up time, 81 (74%) patients in MVP experienced the composite outcome (34 death, 49 ESKD, 71 30% decline in eGFR). In BioVU 13 (68%) patients experienced the composite outcome (1 death, 10 ESKD, 12 30% decline in eGFR). In the pooled Model 3, the effect of 1 unit increase in log UACR at 12 months significantly increased the risk of having composite outcome (adjusted hazard ratio [aHR]=1.61; 95% CI: 1.19-2.17; p=0.002) (**Table 3**).

**Table 3.** The effect of UACR change at 12 months on the risk of the clinical meaningful endpoint among non-diabetic proteinuric patients with 2 APOL1 high risk variants.

|  | BioVU |  | MVP |  | Pooled |  |
| --- | --- | --- | --- | --- | --- | --- |
|  | HR [95% CI] | p | HR [95% CI] | p | HR [95% CI] | p |
| **Clinical Composite Outcome** |  |  |  |  |  |  |
| **Primary analysis (baseline UACR ≥ 420 mg/g; 25 ≤ baseline eGFR < 90)** |  |  |  |  |  |  |
| Model 1 | 2.12 [0.58, 7.81] | 0.26 | 1.51 [1.13, 2.01] | 0.005 | 1.53 [1.16, 2.02] | 0.003 |
| Model 2 | 2.33 [0.55, 9.86] | 0.25 | 1.56 [1.15, 2.10] | 0.004 | 1.58 [1.18, 2.12] | 0.002 |
| Model 3 | 2.55 [0.58, 11.19] | 0.21 | 1.58 [1.16, 2.14] | 0.003 | 1.61 [1.19, 2.17] | 0.002 |
| **Sensitivity analysis 1 (baseline UACR ≥ 420 mg/g; 25 ≤ baseline eGFR < 75)** |  |  |  |  |  |  |
| Model 1 | 2.03 [0.51, 8.03] | 0.32 | 1.54 [1.15, 2.05] | 0.004 | 1.55 [1.17, 2.06] | 0.002 |
| Model 2 | 2.89 [0.56, 14.86] | 0.21 | 1.60 [1.18, 2.18] | 0.003 | 1.63 [1.21, 2.21] | 0.002 |
| Model 3 | N/A* |  | 1.63 [1.19, 2.23] | 0.002 | N/A |  |
| **End Stage Kidney Disease** |  |  |  |  |  |  |
| **Primary analysis (baseline UACR ≥ 420 mg/g; 25 ≤ baseline eGFR < 90)** |  |  |  |  |  |  |
| Model 1 | 3.15 [1.10, 9.00] | 0.03 | 1.74 [1.21, 2.48] | 0.003 | 1.85 [1.32, 2.59] | <0.001 |
| Model 2 | 3.18 [1.13, 8.95] | 0.03 | 1.82 [1.25, 2.67] | 0.002 | 1.94 [1.36, 2.76] | <0.001 |
| Model 3 | 3.74 [1.31, 10.70] | 0.01 | 1.82 [1.25, 2.66] | 0.002 | 1.98 [1.38, 2.82] | <0.001 |
| **Sensitivity analysis 1 (baseline UACR ≥ 420 mg/g; 25 ≤ baseline eGFR < 75)** |  |  |  |  |  |  |
| Model 1 | 2.86 [0.99, 8.25] | 0.05 | 1.81 [1.26, 2.58] | 0.001 | 1.89 [1.35, 2.66] | <0.001 |
| Model 2 | 2.83 [1.00, 8.00] | 0.05 | 1.92 [1.31, 2.80] | 0.001 | 2.01 [1.41, 2.86] | <0.001 |
| Model 3 | 3.77 [1.21, 11.76] | 0.02 | 1.92 [1.31, 2.82] | 0.001 | 2.06 [1.43, 2.96] | <0.001 |

Model 1. adjusted for baseline log transformed UACR.
Model 2. adjusted for baseline log transformed UACR, age, and sex
Model 3. adjusted for baseline log transformed UACR, age, sex, and RAASi
Pooled estimates were done using fixed effects meta-analysis.
Kidney composite endpoint defined as sustained ≥ 30% decline in eGFR, ESKD or death.
\*: Not Available, since the model fitting cannot converge to estimate the HR and 95% CI.

According to the Model 3 for the pooled cohort, a 50% reduction of the UACR was associated with a 28% reduction in the kidney composite endpoint of 30% decline in eGFR, ESKD or death (*aHR*=0.72; 95% CI: 0.59-0.88; p=0.002). Furthermore, we estimated that even a smaller reduction of 30% of the UACR from baseline was associated with a 16% reduction in the kidney composite (*aHR*=0.84; 95% CI, 0.76-0.94; p=0.002).

### The effect of UACR change at 12 months on ESKD endpoint

During the follow up time, 49 patients in MVP and 10 patients in BioVU experienced ESKD. In the pooled Model 3, the effect of 1 unit increase in log UACR at 12 months showed aHR of 1.98 [1.38, 2.82] for the ESKD outcome (**Table 3**). We estimated that a 50% reduction of the UACR was associated with a 38% reduction in the risk of ESKD (*aHR*=0.62; 95% CI, 0.49-0.80; p < 0.001). Furthermore, we estimated that even a smaller reduction of 30% of the UACR from baseline was associated with a 22% reduction in the risk of ESKD (*aHR*=0.78; 95% CI, 0.69-0.89; p < 0.001).

Instead of treating log UACR change as a continuous exposure, we also dichotomized it by the median and defined a two-level (upper half vs. lower half) category variable. We used it as an alternative exposure to estimate the risk for those who were in the upper half of the changes of UACR at 12 months in MVP cohort. After adjusting for baseline log transformed UACR, age, sex, and the use of RAASi, for those in the upper half, the risk of experiencing a clinical composite endpoint was about 2.2 times compared to those in the lower half (aHR=2.19; 95% CI: 1.25-3.82; p=0.006); and the risk of experiencing ESKD was about 2.4 times versus those in the lower half (aHR=2.41; 95% CI: 1.25-4.66; p=0.009) (**Figure 3**).

**Figure 3.**
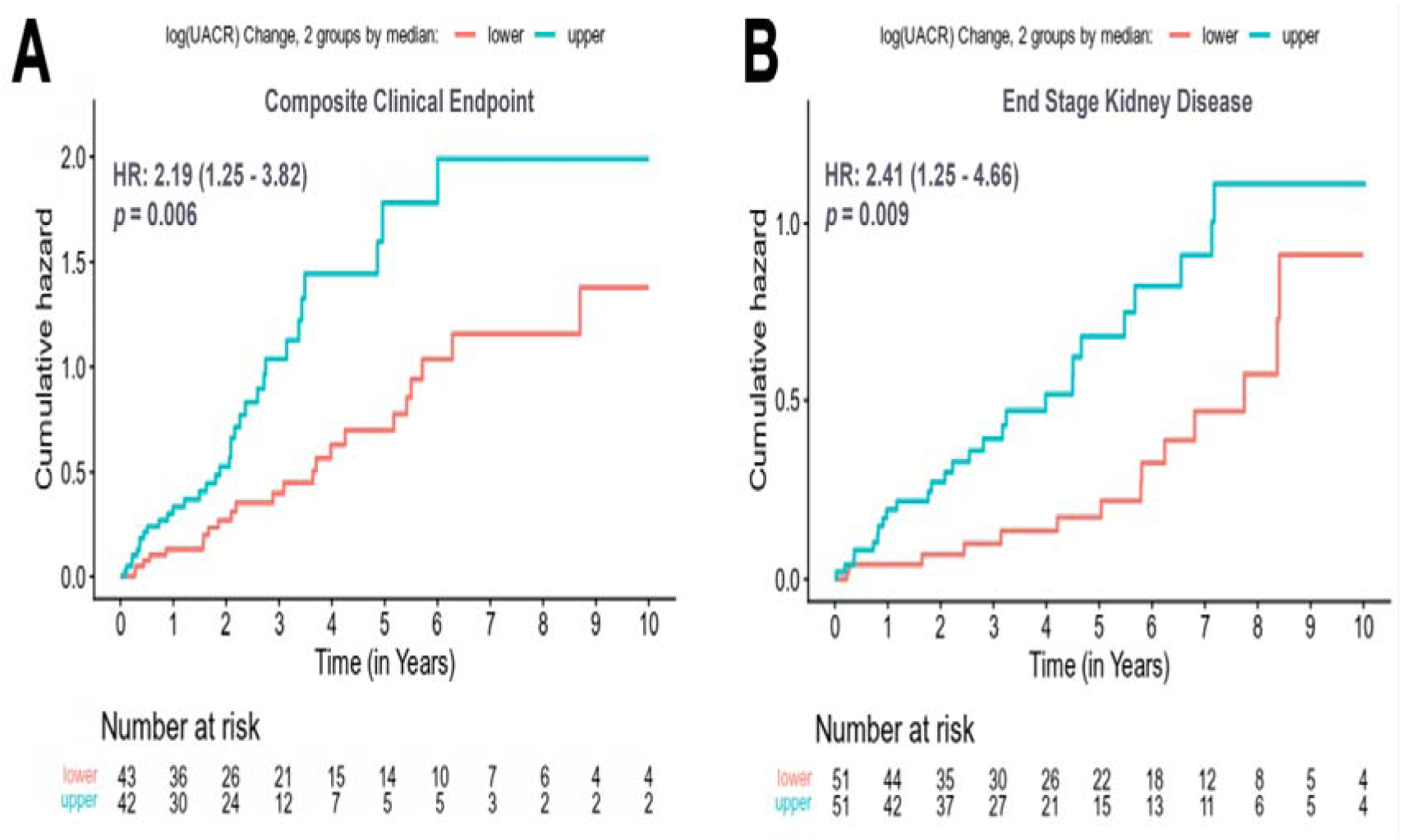
Cumulative Hazard Plot for the composite kidney outcome and ESKD stratified by the dichotomized log UACR change at 12-month in the MVP.

## Sensitivity analysis

We performed sensitivity analyses by evaluating the association of changes in UACR at 12 months and the same primary and secondary outcomes but using the UACR threshold of 321 mg/g (∼0.7 g/g UPCR) for cohort entry (**Supplemental Figure 2 and Supplemental Table 1**). Results were similar and statistically significant, but the effect sizes were smaller for all analyses (**Supplementary Tables 2 and 3**).

## Discussion

Our study in two different predominantly non-FSGS cohorts examined the association of UACR changes at 12 months (reduction and not complete resolution) with the study outcomes in AMKD, showing a strong prognostic association of UACR changes at 12 months and the annual slope of eGFR in 2 years and clinically meaningful endpoints of a 30% decline in eGFR, ESKD and death. These findings support the use of UACR changes as a surrogate endpoint for clinical trials devoted to the treatment of proteinuric AMKD.

Quantitative relationship of changes in proteinuria with kidney function/kidney failure are important to understand what magnitude of reduction in proteinuria will translate into a meaningful reduction in the risk of kidney failure. In our time to event analysis, we estimated that a 50% reduction of UACR was associated with a 28% reduction in the composite outcome of 30% decline in eGFR, ESKD or death. Similarly, a 50% decline was associated with a 38% reduction in the risk of ESRD.

We also explored the effect of UACR reduction of 30% as proposed by the National Kidney Foundation, FDA, and European Medicines Agency in March 2018^13^. In our study, reduction of 30% UACR also significantly predicted a reduction in clinical meaningful endpoints, albeit with a smaller effect size than 50%: 16% and 22% reduction in the risk of the composite outcome and ESKD respectively.

Our findings are consistent with what has been demonstrated in other proteinuric kidney diseases such as FSGS and IgAN, where UACR is now accepted as a surrogate endpoint. Based on the cutoff or threshold identified in this studies of PCR 0.7 g/g, we conducted a sensitivity analysis repeating all our analysis at that threshold, with similar results but with a smaller effect size compared to a threshold of 0.9 g/g, however, with statistical significance showing the ability to predict the reduction of clinical events with UACR changes at 12 months.

These findings are also consistent with prior studies in AMKD populations demonstrating that higher UPCR is associated with increased risk of kidney failure^14^ while reduction in UPCR is associated with improvements in eGFR slope and lower risk of ESKD or death^29^. In a pooled analysis of AASK (n=136) and CRIC (n=180) AMKD cohorts, Rosenberg et al. reported an increased risk of kidney failure (HR: 4.01; 95% CI: 1.96, 8.24) among patients who experienced doubling of UPCR from baseline to three years, irrespective of baseline proteinuria status. Troost et al. evaluated the association of proteinuria change over 26 weeks and subsequent eGFR decline and renal survival among patients with steroid resistant FSGS (n=138; UPCR ≥ 1 g/g ) enrolled in a randomized treatment trial that compared cyclosporine to mycophenolate mofetil plus dexamethasone. A unit reduction in log-UPCR was associated with 3.90 ml/min/1.72m^2^ per year (95% CI: 2.01 to 5.79) improvement in eGFR and substantially lower risk of ESKD or death (HR: 0.23 [95% CI: 0.12 to 0.44]). No significant interactions was observed by APOL1 high-risk status for either eGFR slope (p=0.80) or ESKD or death ( p=0.76) suggesting consistent associations among high risk and low risk strata{Troost, 2021 #37}.

An important aspect to highlight is that our study design aimed to identify the first proteinuric event that met criteria during patient clinical care and that also had eGFR information. This choice, which may seem rather strict, was made to detect proteinuria as early as possible where it may reflect more active disease rather than tubule interstitial scarring, where disease is already non-reversible. Unfortunately, observed proteinuria reflects both, but early detection and targeted intervention is key to increasing the likelihood of success in preventing progression to kidney failure.

The strengths of this study include the inclusion of individuals that had incident proteinuria, supporting that early interventions are key for the reduction in kidney disease progression. Another strength is the inclusion of two very different populations, BioVU include a younger population and MVP and older population with more co-morbidities, despite these differences the effect size was similar on both cohorts. Another strength was the long time of follow-up.

There were also important limitations, first this is a real-world data analysis, and the data has been collected as part of clinical care. Second, the sample size was small given that we aim to incident proteinuria. Our study results are only applicable to proteinuric AMKD and not generalizable to non-proteinuric AMKD. Finally, this is an observational design and subject to unmeasured confounders. Necessitating for this study to be followed by studies that include also prevalent cases and in more advanced stages of the disease and to be replicated in large cohorts.

In summary, APOL1 high risk genotypes are common and present in 12-15% of African Americans in different cohorts and have partially explain the excess risk of CKD progression to ESKD in individuals of African ancestry. Targeted therapeutic development and approvals are desperately needed to treat this large segment of patients with AMKD^30,31^. Our study supports the use of UACR changes at 12 months, as a surrogate endpoint for AMKD. This could facilitate and accelerate the approval of therapeutics currently in clinical trials in a similar way that proteinuria is now an approved surrogate endpoint for FSGS and IgA nephropathy.^31^ Studies testing other potential surrogate outcomes based on eGFR annual slope are needed.

## Data Availability

the data included in this study is not available publicly and can only be obtain with permission from the VA ORD or VUMC institution

## Acknowledgement

We are grateful to the MVP study participants for their contributions to science and all the enrolling sites and the MVP staff and MVP program office (**Supplemental Table 4**). A.M.H. had full access to all the data in the study and assumed responsibility for data integrity. We are also grateful to Vanderbilt University Medical Center for their support to our staff and faculty for their research endeavor. For the BioVU data we are grateful to the AGD/BioVU Pilot project and QC team for the generation of the whole genome data for the AGD linked to Vanderbilt electronic medical center and to the big data team for processing of the synthetic derivative that was used in this project. For BioVU A.M.H. had full access to all the data in the study and assumed responsibility for data integrity. Its contents are solely the responsibility of the authors and do not necessarily represent official views of the National Center for Advancing Translational Sciences or the National Institutes of Health and this publication does not represent the views of the Department of Veteran Affairs or the United States Government.

## Funding acknowledgement

This research is based on data from the Million Veteran Program, Office of Research and Development, Veterans Health Administration, and was supported by MVP000 as well as CSR&D merit award #I01CX001897 titled “Genetic of Kidney Disease and Hypertension in MVP II” (PI: Adriana M. Hung)]. The BioVU project described was supported by CTSA award No. UL1 TR002243 from the National Center for Advancing Translational Sciences. The sequencing of WGS individuals from BioVU, described here, has been funded by the Alliance for Genomic Discovery consisting of NashBio, Illumina and industry partners Amgen, AbbVie, AstraZeneca, Bayer, BMS, GSK, Merck Sharp & Dohme LLC, and Novo Nordisk. The work focus in APOL1 in BioVU was also partially funded by Vertex Pharmaceuticals Incorporated.

## Supplemental material

**Supplemental Figure 1.**
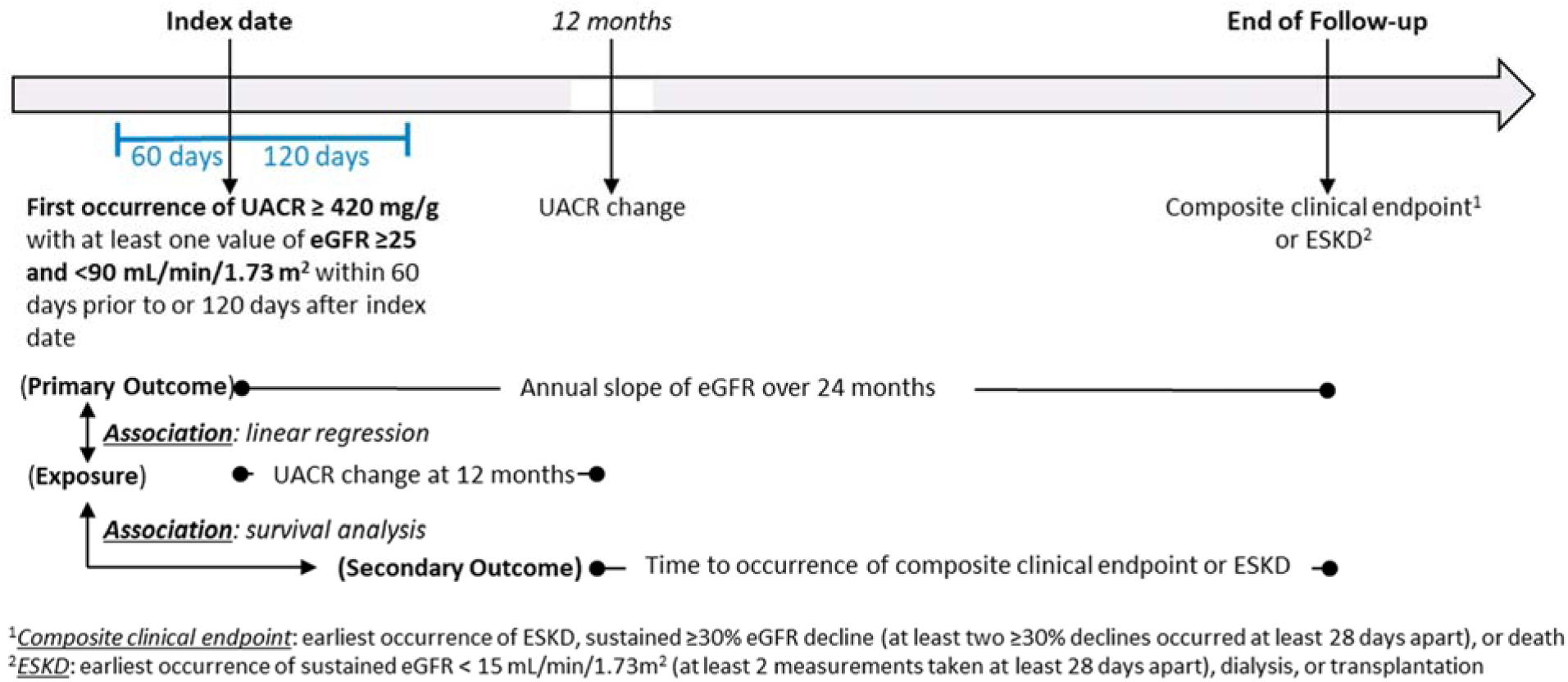
Study design

**Supplemental Figure 2.**
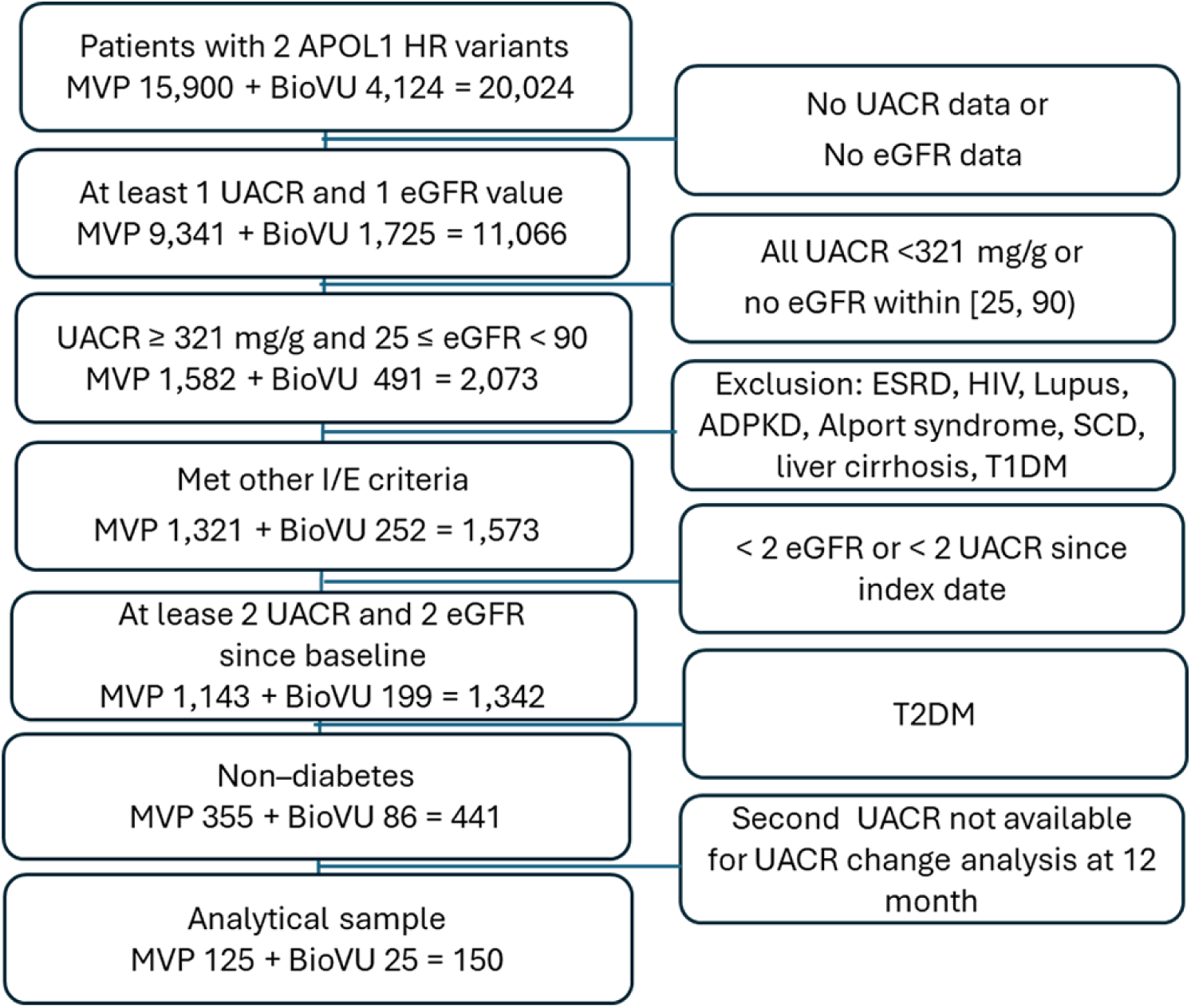
Sensitivity analysis: Flow chart for patients with baseline UACR ≥ 321 mg/g

**Supplemental Table 1.** Clinical characteristics for cohort in sensitivity analysis 2 (i.e., baseline UACR ≥ 321 mg/g, 25 ≤ baseline eGFR0 < 90)

|  | BioVU | MVP | Pooled |
| --- | --- | --- | --- |
| No. patients | 25 | 125 | 150 |
| Age, years (mean, SD) | 47.9 (17.9) | 58.6 (14.8) | 56.8 (15.8) |
| Males (n, %) | 15 (60.0%) | 118 (94.4%) | 133 (88.7%) |
| eGFR, mL/min/1.73m<sup>2</sup> (mean, SD) | 48.1 (19.0) | 46.7 (16.6) | 46.9 (17.0) |
| UACR, mg/g (mean, SD) | 1275.9 (1388.2) | 997.8 (1014.7) | 1044.2 (1085.4) |
| ACEi or ARBs (n, %) | 10 (40.0%) | 94 (75.2%) | 104 (69.3%) |
| SGLT2 (n, %) | 0 (0.0%) | 1 (0.8%) | 1 (0.7%) |
| FSGS, baseline (n, %) | 3 (12.0%) | 0 (0.0%) | 3 (2.0%) |
| FSGS, within follow up (n, %) | 3 (12.0%) | 7 (5.6%) | 10 (6.7%) |
| Follow-up, yrs (mean, SD) | 3.7 (4.4) | 3.5 (3.6) | 3.5 (3.7) |
| Follow-up, yrs (median, IQR) | 1.8 [0.8;5.5] | 2.6 [1.1;4.3] | N/A* |
| Annual eGFR slope over 24 months (mL/min/1.73m<sup>2</sup>/year), 95% CI | -6.56 [-13.18, 0.06] | -3.60 [-5.03, -2.17] | -3.73 [-5.13, -2.33] |
| Geometric mean percentage change in proteinuria at 12 months (%), 95% CI | -51.1% [-70.6%, -18.7%] | -52.7% [-61.5%, -41.9%] | -52.5% [-60.6%, -42.6%] |
| **Meaningful clinical endpoints** |  |  |  |
| 30% decline in eGFR | 14 (56.0%) | 78 (62.4%) | 92 (61.3%) |
| End stage kidney disease | 12 (48.0%) | 52 (41.6%) | 64 (42.7%) |
| death | 3 (12.0%) | 39 (31.2%) | 42 (28.0%) |
| Composite | 15 (60.0%) | 88 (70.4%) | 103 (68.7%) |

N/A\*: Not Available, since there is no feasible approach to get pooled median IQR without combining individual data.

**Supplemental Table 2.** Association between log-UACR changes from baseline to 12 months and eGFR slope over 24 months among non-diabetic patients with 2 APOL1 high risk variants for patients with baseline UACR ≥ 321 mg/g.

|  | **BioVU** |  | **MVP** |  | **Pooled** |  |
| --- | --- | --- | --- | --- | --- | --- |
|  | *b* [95% CI] | *p* | *b* [95% CI] | *p* | *b* [95% CI] | *p* |
| ***Sensitivity analysis (baseline UACR $\geq 321$ mg/g; $25 \leq$ baseline eGFR < 90)*** |  |  |  |  |  |  |
| Model 1 | -1.38 [-5.96, 3.20] | 0.54 | -1.23 [-2.10, -0.37] | 0.005 | -1.24 [-2.08, -0.40] | 0.004 |
| Model 2 | -1.75 [-6.62, 3.12] | 0.46 | -1.31 [-2.18, -0.44] | 0.003 | -1.32 [-2.17, -0.48] | 0.002 |
| Model 3 | -2.88 [-8.15, 2.39] | 0.27 | -1.31 [-2.19, -0.43] | 0.004 | -1.36 [-2.22, -0.50] | 0.002 |
| ***Sensitivity analysis (baseline UACR $\geq 321$ mg/g; $25 \leq$ baseline eGFR < 75)*** |  |  |  |  |  |  |
| Model 1 | -1.44 [-6.12, 3.24] | 0.53 | -1.56 [-2.39, -0.73] | <0.001 | -1.56 [-2.36, -0.75] | <0.001 |
| Model 2 | -1.86 [-6.86, 3.13] | 0.44 | -1.60 [-2.43, -0.78] | <0.001 | -1.61 [-2.42, -0.81] | <0.001 |
| Model 3 | -2.04 [-7.07, 2.98] | 0.40 | -1.63 [-2.46, -0.79] | <0.001 | -1.64 [-2.45, -0.83] | <0.001 |

**Supplemental Table 3.** The effect of UACR change at 12 months on the risk of the clinical meaningful endpoint among non-diabetic proteinuric patients with 2 APOL1 high risk variants.

|  | **BioVU** |  | **MVP** |  | **Pooled** |  |
| --- | --- | --- | --- | --- | --- | --- |
|  | *HR [95% CI]* | *p* | *HR [95% CI]* | *p* | *HR [95% CI]* | *p* |
| ***Clinical Composite Outcome*** |  |  |  |  |  |  |
| ***Sensitivity analysis (baseline UACR ≥ 321 mg/g; 25 ≤ baseline eGFR < 90)*** |  |  |  |  |  |  |
| Model 1 | 1.94 [0.91, 4.16] | 0.09 | 1.39 [1.08, 1.78] | 0.01 | 1.43 [1.13, 1.81] | 0.003 |
| Model 2 | 1.91 [0.86, 4.20] | 0.11 | 1.41 [1.09, 1.82] | 0.008 | 1.45 [1.14, 1.85] | 0.003 |
| Model 3 | 1.63 [0.72, 3.68] | 0.24 | 1.41 [1.09, 1.83] | 0.009 | 1.43 [1.12, 1.83] | 0.005 |
| ***Sensitivity analysis (baseline UACR ≥ 321 mg/g; 25 ≤ baseline eGFR < 75)*** |  |  |  |  |  |  |
| Model 1 | 1.86 [0.85, 4.06] | 0.12 | 1.40 [1.08, 1.81] | 0.01 | 1.44 [1.12, 1.83] | 0.004 |
| Model 2 | 1.71 [0.77, 3.79] | 0.19 | 1.43 [1.09, 1.86] | 0.009 | 1.45 [1.13, 1.87] | 0.004 |
| Model 3 | 1.89 [0.82, 4.36] | 0.14 | 1.44 [1.10, 1.88] | 0.009 | 1.47 [1.14, 1.91] | 0.003 |
| ***End Stage Kidney Disease*** |  |  |  |  |  |  |
| ***Sensitivity analysis (baseline UACR ≥ 321 mg/g; 25 ≤ baseline eGFR < 90)*** |  |  |  |  |  |  |
| Model 1 | 3.23 [1.30, 8.02] | 0.01 | 1.68 [1.21, 2.32] | 0.002 | 1.81 [1.33, 2.45] | <0.001 |
| Model 2 | 4.23 [1.16, 15.43] | 0.03 | 1.71 [1.22, 2.38] | 0.002 | 1.81 [1.31, 2.49] | <0.001 |
| Model 3 | 4.88 [1.10, 21.63] | 0.04 | 1.71 [1.22, 2.40] | 0.002 | 1.80 [1.29, 2.51] | <0.001 |
| ***Sensitivity analysis (baseline UACR ≥ 321 mg/g; 25 ≤ baseline eGFR < 75)*** |  |  |  |  |  |  |
| Model 1 | 3.15 [1.29, 7.69] | 0.01 | 1.73 [1.24, 2.40] | 0.001 | 1.85 [1.36, 2.52] | <0.001 |
| Model 2 | 4.91 [1.19, 20.20] | 0.03 | 1.78 [1.27, 2.51] | 0.001 | 1.89 [1.35, 2.63] | <0.001 |
| Model 3 | 5.21 [1.22, 22.26] | 0.03 | 1.80 [1.27, 2.54] | 0.001 | 1.90 [1.36, 2.66] | <0.001 |

N/A\*: Not Available, since the model fitting cannot converge to estimate the HR and 95% CI.

**Supplemental Table 4.**
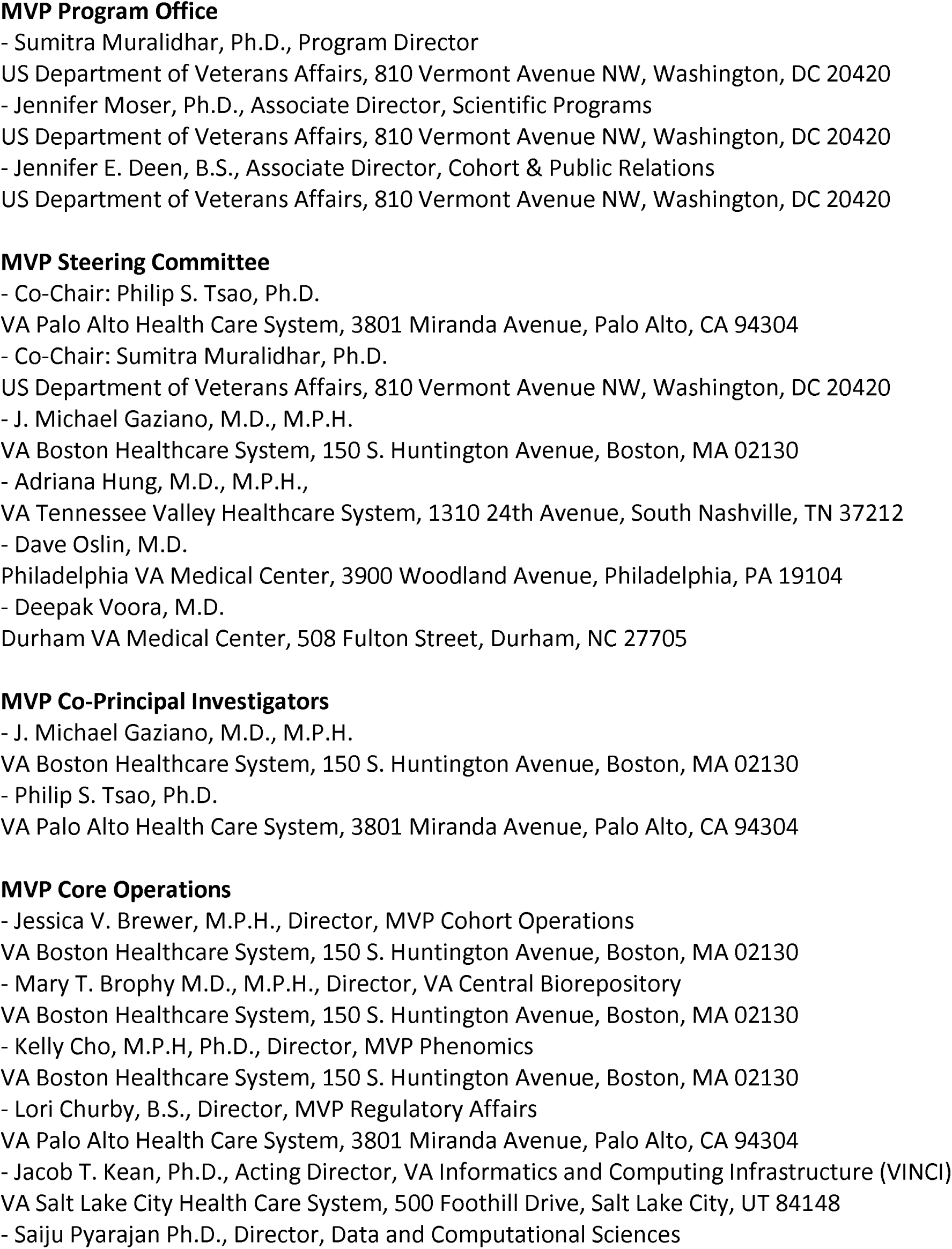
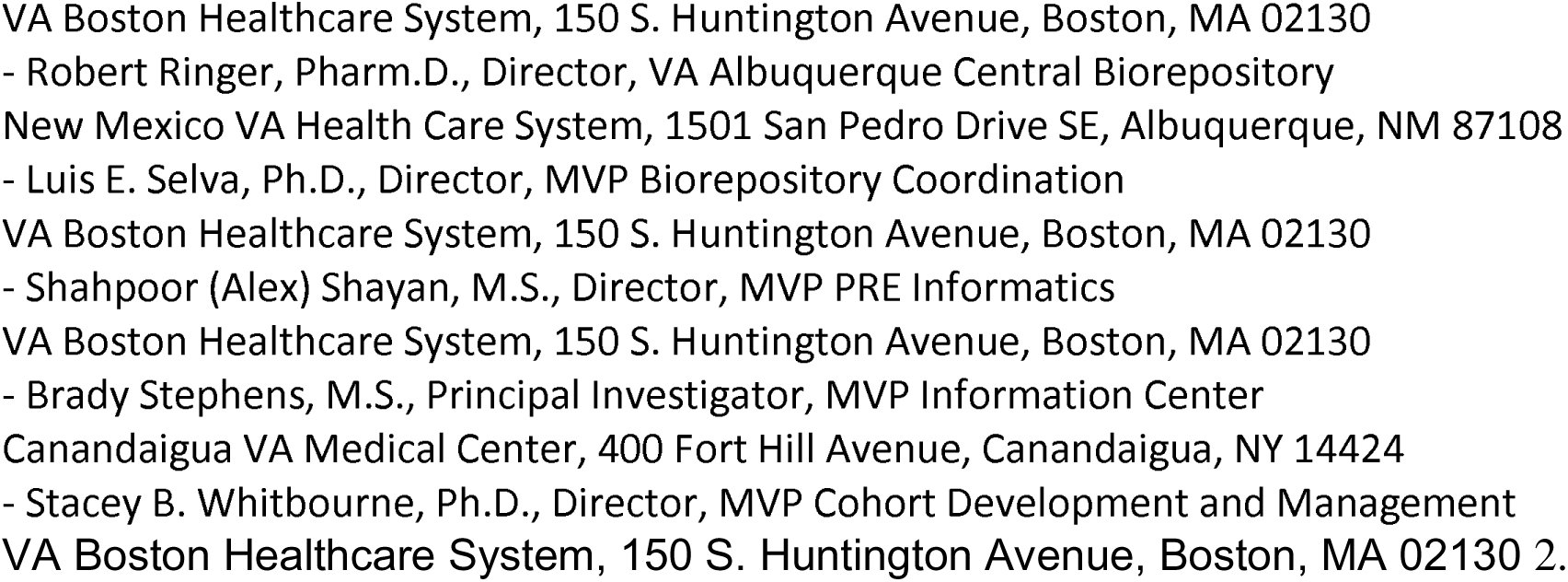
MVP Core Acknowledgements for Publications_October 2025.

